# Causes, Predictors, and Costs of Unplanned 30-Day Readmissions in Patients with Right Heart Failure

**DOI:** 10.1101/2021.06.28.21257859

**Authors:** Amod Amritphale, Jesan Zaman, Saad Awan, Nupur Amritphale, G. Mustafa Awan, Suvo Chatterjee, Bassam Omar, Gregg C. Fonarow

**Author notes:** Co-first authors; contributed equally to all aspects of the research (Amod Amritphale, Jesan Zaman). Disclaimers: GCF discloses consulting for Abbott, Amgen, AstraZeneca, Bayer, Edwards, Janssen, Medtronic, Merck, and Novartis. Sources of support: None.

## Abstract

**OBJECTIVES:** The aim of this study was to describe the frequency, causes, factors, and costs associated with right heart failure encounters.

**BACKGROUND:** Multiple studies have looked into heart failure readmissions but there is no study to our knowledge specifically analyzing patients with right heart failure.

**METHODS:** Patients with right heart failure diagnosis were identified using the 2017 Nationwide Readmission Database (NRD) and were evaluated for the rates, predictors, and costs of unplanned 30 days readmission. Weighted analysis was performed to obtain nationally representative data.

**RESULTS:** We identified 7142 patients with right heart failure of whom 21% had an unplanned readmission within 30 days of the index discharge. Patients with history of Coronary artery bypass grafting (*p*=0.033; OR 2.359; 95% CI 1.071 - 5.197), Chronic kidney disease (p<0.001; OR 1.607; 95% CI 1.402 - 1.843), atrial fibrillation (*p*=0.014; OR 1.417; 95% CI 1.072–1.873) had high odds of unplanned 30 day readmissions while obesity (p<0.001; OR 0.686; 95% CI 0.594 - 0.792) had negative odds of such readmissions.

**CONCLUSIONS:** Thirty-day unplanned readmissions remain a significant issue affecting over 1 in 5 patients with right heart failure. Early unplanned readmissions are associated with significant mortality and financial burden in these patients.

## INTRODUCTION

Unplanned readmissions over a 30-day period after index hospitalization are known to occur in up to 20% of Medicare beneficiaries with heart failure (HF) being a major driver of readmissions and hence penalties. Heart failure is one of the most frequent diagnosis for 30-day readmissions^1^, and as projected for 2030, financially, will have an increase of 215% in direct costs, 80% in indirect costs, and 25% in prevalence respectively^2^.

Unfortunately, despite the fact that right heart failure (RHF) in the presence of left-sided failure (LVF) is the most common scenario, analysis of patients with right heart failure remains incompletely elucidated.

Development of right heart failure has been hypothesized to be secondary to multitude of mechanisms including ischemia, myocarditis, development of pulmonary hypertension, severe LVF resulting in decreased coronary perfusion for the right ventricle and codependence leading to impair RV diastolic function by increasing pericardial constraint. With improving medical therapy and guideline adherence, patients have improved survival, RV failure may constitute a “common final pathway” in the progression of congestive heart failure^3^.

To the best of our knowledge, there is no comprehensive synopsis looking specifically into the inpatient mortality using a nationally representative data in the International Classification of Diseases (ICD)-10th Revision era. The specificity of diagnostic and procedure codes have immensely improved with the advent of ICD-10 codes which can give us better insight into the causes and associations of inpatient admissions and mortality.^4^ We utilized the data made available by the Healthcare Cost and Utilization Project (HCUP) which was created through a Federal-State-Industry partnership and sponsored by the Agency for Healthcare Research and Quality (AHRQ) and it includes the largest collection of longitudinal hospital care data in the United States, with all-payer, encounter-level information. We followed the direction of previous well conceived studies using the database that have evaluated the risks of readmissions in patients with carotid stenting and pediatric sepsis^34,35^.

In this study, we present a comprehensive all-cause, all-comer and all-payer analysis of inpatient data extracted from the HCUP Nationwide Readmission Database (NRD) using over 35 million inpatient encounters in the United States.

## Methods

The NRD is a nationally representative sample of all-age, all-payer discharges from U.S. nonfederal hospitals produced by the Healthcare Cost and Utilization Project of the Agency for Healthcare Research and Quality. This database is composed of discharge-level hospitalization data from 28 geographically dispersed states across the United States of America. Approximately, it has 18 million discharges for the year 2017 (Weighted estimated to roughly 36 million discharges). The dataset used in the present study represents 60% of the U.S. population and 58.2% of all U.S. hospitalizations. We have presented a flowchart in Figure 1 that provides details for included/excluded patient encounters with both weighted and unweighted numbers.

Within a calendar year, hospitalizations and rehospitalizations can be determined using a de-identified unique patient linkage number assigned to each patient, which enables tracking of patients across hospitals within a state. Individual patients in the NRD are assigned up-to 40 diagnosis codes and 25 procedure codes for each hospitalization. The study was approved for an exempt status by the Institutional Review Board at the University of South Alabama.

We defined right heart failure with the procedure codes from ICD-10 as in Appendix Table 1. The primary outcome was first readmission after being diagnosed with right heart failure during the year 2017. If a patient had multiple right heart failure admissions in a year, only the first encounter where the patient had right heart failure was used as index admissions for the analysis. Patients who died during their initial hospitalizations were excluded. The cohort patients admitted in the month of December for the index admission were also excluded, as they may not have 30 days of follow-up, leading to immortal time bias.

Diagnosis a lot of the times get established early based on initial symptoms and then the overall main diagnosis of right heart failure is subsequently discovered during the actual course of admission. The initial diagnosis cannot be assumed and thus up to 40 diagnosis codes were utilized in our study. Patient information is also not carried on from one year to the next, and so patients cannot be tracked across years. Elective admissions were not counted as unplanned readmissions. We also wanted to ensure that regarding readmission it was not just a short 1-night admission to ensure follow up, and this is taken care of because the NRD only captures patients that have inpatient status, not observation which is less than 24hrs.

ICD-10 codes were used to define clinical variables and chosen based on clinical importance. The All Patient Refined-Diagnosis Related Group (APR DRG) Mortality risk score and APR DRG Severity of Illness calculations were also performed in the analysis.

Additional data on hospital bed size, control/ownership of hospital, hospital urban rural designation, and teaching hospitalwere analyzed. Cost analysis was performed by analyzing data on primary expected payer, quartile of median household income, index admission length of stay (days), index admission cost (U.S.$), discharge destination to home/self-care, transfer to other hospital, care home, discharge against medical, advice, readmission length of stay (days), readmission charges & costs (U.S.$), and readmission death. The costs were determined by multiplying the hospital charges with the Agency for Healthcare Research and Quality’s all-payer cost-to-charge ratios for each hospital. The causes of readmission were determined by the first diagnosis on the basis of Clinical Classification Software codes (CCSR). All ICD-10 and CCSR codes used in this study have been presented in the appendix section.

Statistical analysis was performed using IBM SPSS Statistics for Windows, version 1.0.0.118 (IBM Corp., Armonk, N.Y., USA) using 2-sided tests and a significance level of 0.05. Multiple logistic regressions were used to determine the predictors of readmission within each time period, with adjustments for all variables except the elective variable.

## Results

### Population Characteristics and Descriptive Results

The NRD included 30,413,839 inpatient encounters from January through December 2017 among hospitalized patients 18 years of age and older. Using weighted analysis, we identified 7142 individual patients who were diagnosed with right heart failure (figure 1). After index admission with right heart failure, 21% patients (n=1501) had an unplanned 30-day readmission.Table 1 gives a detailed synopsis of the baseline and comorbidities associated with early unplanned readmission.

### Cost analysis and length of stay results

The costs were determined by multiplying the hospital charges with the Agency for Healthcare Research and Quality’s all-payer cost-to-charge ratios for each hospital. The median hospital costs of index encounters were $12881 [7439 – 23857]. The cost for the readmission encounters was $11522 [IQR $6726 – $21591]. The median length of stay for index admission without readmission and readmission encounters were 5 days (IQR 3-9 days) and 6 days (IQR 3-10 days) (*P*=0.037) respectively.

### Predictors of unplanned readmissions

Table 2 presents the results of multivariable logistic regression analysis for predictors of unplanned 30 day readmissions for right heart failure. Patients with history of Coronary artery bypass grafting (*p*=0.033; OR 2.359; 95% CI 1.071 - 5.197), Chronic kidney disease (p<0.001; OR 1.607; 95% CI 1.402 - 1.843), atrial fibrillation (*p*=0.014; OR 1.417; 95% CI 1.072 – 1.873) had high odds of unplanned 30 day readmissions while obesity (p<0.001; OR 0.686; 95% CI 0.594 - 0.792) had negative odds of such readmissions. Forrest plot analysis of comorbidities associated with 30 days readmission is presented (Figure 2). Causes and frequencies of primary diagnosis category for readmission encounters based on Clinical Classification Software refined (CCSR) categories are presented in Figure 3.

## Discussion

The most common cause of readmission for Medicare patients in the United States, with 30 day readmission rates more than 10% is chronic heart failure^4^. CHF causes Americans nearly 30.7 billion dollars a year, which represents a significant financial burden on our healthcare industry^5^. A study from the American Heart Association forecasted costs to increase as high as $53 billion by 2030^6^. With costs like that, it is quite obvious why there is so much attention in reducing the overall burden, with a special focus on readmissions. The hospital readmissions reduction program, which came into action on October 1st, 2012, aimed to improve communication and care coordination to better engage patient and caregivers in discharge plans in an attempt to reduce avoidable readmissions^7^. There were various conditions that CMS specifically looked at in regards to their program, one of them being heart failure. Right heart failure and its implications in CHF have not been evaluated extensively as many views it as secondary to left side heart failure in the prognosis of CHF. In an effort to change that and initiate a discussion, this study is the first comprehensive readmission analysis of patients with right heart failure. This is closest to and represents the real-life practice patterns as it inculcates the inherent benefit of utilizing highly specific ICD-10 codes as a baseline. It also represents data based on the most recent guidelines as it takes a few years to inculcate guidelines in everyday practice.

There have been numerous studies that showed readmission rates within 30 days being associated with poor outcomes^8^. Most studies describe heart failure in general, such as the one by Dr. Greene et al, who described how although recent trends showed reduction in hospital duration stays and a decrease in, in-hospital and 30 day mortality, there has been an increase in 30 day readmission rates and discharges to skilled nursing facilities^9^. This same study showed that among Medicare patients that were admitted for heart failure from 2008 to 2010, approximately 67% more readmitted and almost 36% died within one year of that index admission^9^. There can be many factors attributed to these alarming statistics, one of them in particular (the CHAMP-HF registry) detailed a gap in standardized medical therapy, showing that among patients who were eligible, almost 27% were not prescribed an ACE inhibitor which has showed to uniquely reverse cardiovascular remodeling in heart failure and improve ejection fraction^10,11^. Essentially there is plenty of data on heart failure in general, but not enough on specifically right heart failure, and that is the area we decided to address in our study.

Within our study more than one out of every six patients had an unplanned readmission within just 30 days of the index discharge. In the CHARITEM registry, right heart failure accounted for 2.2% of all heart failure admissions and was secondary to LV failure in more than 20% of cases, this percentage extrapolated to a larger population is rather significant^12,13^. One common point that most people know about right heart failure is that the most common etiology of it is left heart failure. The interesting point about this, is that patients with both left and right heart failure only have a 23% two year survival rate, this percentage jumps all the way up to 71% if the patient only has left ventricular failure alone^11,14,15^.

Utilizing data from 2017 NRD And the newer ICD 10 codes, the 30 day re admission rate from right heart failure was 17.9%, This was rather close to the rates after primary diagnosis of pneumonia (18.69%), cardiac dysrhythmias (17.27%), and UTI (18.66%) found any study reported in the 2014 NRD; Interestingly enough, that same study also reported that the 30-day readmission rate after the primary diagnosis of congestive heart failure was 25.70%, however there was no measure for right heart failure specifically^16^. This highlights the issue that we are discussing regarding the lack of proper investigation into right heart failure specifically, and its’ readmission rates.

Right heart failure is essentially a clinical diagnosis and it is usually a resultant of left ventricular dysfunction^17^. A study over 2 years showed a 45% mortality of patients with right ventricular dysfunction compared to 7% of those without^18^. This highlights the importance of early diagnosis and intervention so that we can improve the overall health outcomes of our patient population.

We found that the most common causes of readmission for patients with RV failure diagnosis on the first admission were heart failure, followed by septicemia and respiratory failure; insufficiency; arrest. The study refers to causes of readmissions in patients with an initial diagnosis of right heart failure. Being able to properly manage these causes of readmission on initial admission is crucial to preventing a rather quick return to the hospital. Improving readmission rates for heart failure has been studied many times, some ways that showed productive results was to schedule appropriate follow up, providing one to one in patient education, making follow up calls within one to two days of discharge, and then again within one to two weeks of discharge, and finally employing the teach back approach^9^. The issue of sepsis is also of concern as it can speed up cardiovascular dysfunction that was existing prior to it^19^. The link between heart failure and sepsis is so prominent that a group of researchers are working on utilizing a risk assessment tool incorporating independent predictors of sepsis death, including advanced age, chronic lung disease, higher platelet counts, lowers vitamin D levels, and male sex^20^. As mentioned, the most common etiologies of right ventricular failure is either existing or new onset cardiac or pulmonary disease. Failure due to lung disease is well known to be called cor pulmonale^21^. The range of pulmonary disease linked with failure from lung disease is extremely broad. That being said, there is an interesting study that detailed the many advancements in the management of acute respiratory distress syndrome otherwise known as ARDS. This study talked about important strategies such as lung protective ventilation and fluid conservative therapy as well as other techniques used for management. However, despite all these advancements, mortality from ARDS is still high, and one of the biggest reasons of this is in fact acute cor pulmonale^22^. All of this is to simply highlight the importance of paying attention to right ventricular failure as a unique pathogenetic process that must be managed cohesively with associated risk factors. Prior studies have shown that patients with advanced coronary disease, especially those dependent on mechanical therapies or having multiple coronary stents have poor outcomes^36,37^.

Our data also showed that prior coronary bypass graft had the highest odds ratio (p=0.02, OR 2.52, CI 1.15-5.52) and in our admission analysis as shown in the Forrest weaver plot table X. A study detailed how this procedure is the most common cardiac surgery performed in the entire world with almost 200,000 cases in the US alone^23,24^. The way to look at this is actually a bit different, because the way decrease the amount of coronary artery bypass grafts performed is quite simply by controlling the risk factors that lead to coronary artery disease itself. Some of these risk factors of course include age, gender, hypertension, hyperlipidemia, diabetes and so forth^25^. An interesting note that while obesity is commonly known to be associated with many comorbidities, In our study it actually showed to have a negative odds of readmission at 0.753 (P<0.001, 0.648-0.876).

Being able to evaluate all of these factors is absolutely crucial in controlling the overabundant health care cost in the United States as well as improving the overall quality of care and quality of life of our patients. The length of days that a patient spends in the hospital on the initial admission was about 5 (the median of index admission length of days) according to our data, while the duration of days on a readmission was one extra (6 days, median of index admission length of days). Hence, we are spending more money during the encounter of the readmission then we did even on the initial one; the difference actually comes out to a little more than $1000. Once again although this may seem like a small number, when extrapolated to the entire country it creates a big dent in our healthcare costs. Medicare accounted for about 68.2% of our entire patient group which places a large financial strain on an already struggling system. The United States spend for medical care almost twice as much as other high income countries around the world as detailed in many studies such as by Papanicolas et al^26^. Further research is needed to identify the subgroup of patients with right heart failure who will be safe for early hospital discharge as has been done for other conditions like for acute myocardial infarction^38^.

Being able to control the various risk factors and paying more attention to right ventricular failure itself will help optimize care and is absolutely critical to the infrastructure over medical system.

The risk of mortality on re admission is also of great concern. We analyzed the APR DRG mortality risk and severity of illness (as shown in Table X) in our study and found that patients who are re admitted within 30 days have approximately 6% increase in the major likelihood of dying as well as a 4% increase in the major loss of function. All in all, this suggests that careful decision about creating a plan by multi-disciplinary team that includes close follow up with a primary care physician but that also includes infectious disease specialist, echo cardiologists, and cardiac surgeons is crucial for making positive steps towards reducing readmissions and healthcare costs after discharge. Another interesting anecdote from the DAPA-HF trial, was the emergence of drugs such as dapagliflozin that have shown to have significant reduction in death due to cardiovascular causes and hospitalization in heart failure^27^.

Adjusting healthcare to the current times of the world is always crucial, and with the increased utilization of phone calls and telecommunicating and or monitoring due to COVID, it may be beneficial to re-analyze a previous study that did not show any benefit, and attempt to acquire more data to see if the use of telecommunication is beneficial to patients^28^.

The HRRP had good intentions with its’ aim to reduce 30 day readmission rates, but the financial implications of the program had numerous negative consequences with some studies showing an actual increase in both short term and long term mortality after a heart failure admission. To say that the program needs to be revamped and adjusted would be an understatement, especially considering that there was not much of a reduction in readmissions after its inception^29,30^.

Our study is the first of its kind to demonstrate the importance of evaluating various characteristics, demographics, and comorbidities in order to reduce the risk and likelihood of re admissions within 30 days of an initial discharge. The median index admission hospital charge on readmission was approximately $46,000 (compared to that of approx. $44,000 from an initial admission). Our results are very similar to that of previous studies such as from Timothy Welch et al^31^. This is actually significantly more than the costs of heart failure readmissions^32^. This highlights the importance of early, detailed and close interventions to prevent the re admission of these patients, especially with those who have several risk factors for re admission. With all of this knowledge we should have a more focused and individualized follow-up intervention for each patient that we have, as opposed to normal broad stroke interventions that are in place currently^33^. More work is necessary to increase awareness about the right heart failure and factors associated with worse outcomes using social media, YouTube and Smart Phone applications as have been done for other disease conditions in the past^39,40^. Further research should follow on the lines of using artificial intelligence and the deep neural network to identify unidentified associations as has been pioneered in published studies on carotid artery stenting and Covid-19^34,41^.

### Study limitations

By design, NRD does not allow the determination of regional variations within the dataset. This study does not capture data of patients who were admitted as an outpatient observation status and represents only hospital admissions. Other limitations include inaccurate coding or subjective coding by physicians as well as patients admitted outside the tracking hospitals could have been missed. As with any observational data, the results do not suggest causal relationships as there can be other unmeasured confounders. Diagnosis codes are assigned to patients throughout an admission process based on various elements such as symptoms, labs and procedures ordered, and the NRD does not detail on validation studies to confirm the diagnosis codes used during inpatient admissions.

## Conclusion

Our analysis suggests that over 1 in 5 patients admitted with right heart failure end up with an unplanned readmission within 30 days of discharge. There are several areas of improvement in the quality of care the patients with right heart failure receive. This study will pave the way for a more comprehensive approach for identification and may help develop protocols to help this subsection of population at risk for one of the highest incidence of unplanned readmissions.

## Data Availability

All data is publicly available, we utilized the NRD Database

## Acknowledgment

The authors are grateful to the Healthcare Cost and Utilization Project for providing the data used in the analysis.

## Abbreviations

NRD: Nationwide Readmission Database
ICD: International Classification of Diseases
CMS: Centers for Medicare & Medicaid Services
RSRR: Risk-Standardized Readmission Rates
HRRP: Hospital Readmissions Reduction Program
CAD: coronary artery disease
CVA: Cerebro- Vascular accidents
CABG: Coronary artery bypass grafting
GI: Gastrointestinal
COPD: Chronic obstructive pulmonary disease
CKD: Chronic kidney disease
AKI: Acute kidney injury
CCSR: Clinical Classification Software refined
IQR: Inter Quartile Range
APR-DRG: All Patient Refined-Diagnosis Related Group
MACCE: Major Adverse Cardiovascular and Cerebrovascular Events
PAD: Peripheral artery disease

## Notes

### Competing Interest Statement

The authors have declared no competing interest.

### Clinical Trial

No Clinical trials were conducted

### Funding Statement

No Funding

### Author Declarations

The study is based on publicly available deidentified database and was approved for exempt status by the IRB at University of South Alabama

## Citations

1. Fida, N, Pina, IL. Trends in heart failure hospitalizations. Curr Heart Fail Rep 2012; 9: 346–353.

2. Heidenreich, PA, Sahay, A, Kapoor, JR. Divergent trends in survival and readmission following a hospitalization for heart failure in the Veterans Affairs health care system 2002 to 2006. J Am Coll Cardiol 2010; 56: 362–368.

3. Voelkl N. F., Quaife R. A., Leinwand L. A., Barst R. J., McGoon M. D., Meldrum D. R., Dupuis J., Long C. S., Rubin L. J., Smart F. W., Suzuki Y. J., Gladwin M., Denholm E. M., & Gail D. B. National Heart, Lung, and Blood Institute Working Group on Cellular and Molecular Mechanisms of Right Heart Failure. Right ventricular function and failure. Report of the National Heart, Lung, and Blood Institute working group on cellular and molecular mechanisms of right heart failure. Circulation. 2006 Oct. 24 114 (17): 1883–91.

4. Medicare Payment Advisory Commission. (2007, June). Payment policy for inpatient readmissions. In Report to the Congress: promoting greater efficiency in Medicare (pp. 102–120). Washington, DC: MedPAC.

5. Heidenreich PA, Albert NM, Allen LA, et al. Forecasting the impact of heart failure in the United States: a policy statement from the American Heart Association. Circ Heart Fail. 2013;6(3):606–619. doi:10.1161/HHF.0b013e318291329a

6. https://www.cms.gov/Medicare/Medicare-Fee-for-Service-Payment/AcuteInpatientPPS/Readmissions-Reduction-Program as accessed on 10/02/20

7. https://www.aha.org/factsheet/2016-01-18-aha-fact-sheet-hospital-readmissions-reduction-program as accessed on 10/02/20

8. Arundel C, Lam PH, Khosla R, et al. Association of 30-Day All-Cause Readmission with Long-Term Outcomes in Hospitalized Older Medicare Beneficiaries with Heart Failure. Am J Med. 2016;129(11):1178–1184. doi:10.1016/j.amjmed.2016.06.018

9. Ziaeian B, Fonarow GC. The Prevention of Hospital Readmissions in Heart Failure. Prog Cardiovasc Dis. 2016;58(4):379–385. doi:10.1016/j.pcad.2015.09.004

10. Greene SJ, Butler J, Albert NM, et al. Medical Therapy for Heart Failure With Reduced Ejection Fraction: The CHAMP-HF Registry. J Am Coll Cardiol. 2018;72(4):351–366. doi:10.1016/j.jacc.2018.04.070

11. Peri-Okonny PA, Mi X, Khariton Y, et al. Target Doses of Heart Failure Medical Therapy and Blood Pressure: Insights From the CHAMP-HF Registry. JACC Heart Fail. 2019;7(4):350–358. doi:10.1016/j.jchf.2018.11.011

12. Jackson SL, Tong X, King RJ, Loustalot F, Hong Y, Ritchey MD. National Burden of Heart Failure Events in the United States, 2006 to 2014. Circ Heart Fail. 2018;11(12):e004873. doi:10.1161/CIRCHEARTFAILURE.117.004873

13. Searle J, Frick J, Möckel M. Acute heart failure facts and numbers: acute heart failure populations. ESC Heart Fail. 2016;3(2):65–70. doi:10.1002/ehf2.12092

14. Mandras SA, Desai S. Right Heart Failure. [Updated 2020 Jul 21]. In: StatPearls [Internet]. Treasure Island (FL): StatPearls Publishing; 2020 Jan-. Available from: https://www.ncbi.nlm.nih.gov/books/NBK459381/ as accessed on 9/24/20

15. Ziaeian B, Fonarow GC. Epidemiology and aetiology of heart failure. Nat Rev Cardiol. 2016 Jun;13(6):368–78. doi: 10.1038/nrcardio.2016.25. Epub 2016 Mar 3. PMID: 26935038; PMCID: PMC4868779.

16. US Agency for Healthcare Research and Quality. Introduction to the HCUP Nationwide Readmissions Database (NRD) 2014. https://www.hcup-us.ahrq.gov/db/nation/nrd/NRD_Introduction_2014.jsp. Accessed 9/24/20

17. Mehra MR, Park MH, Landzberg MJ, Lala A, Waxman AB; International Right Heart Failure Foundation Scientific Working Group. Right heart failure: toward a common language. J Heart Lung Transplant. 2014; 33:123–126. doi: 10.1016/j.healun.2013.10.015.

18. Melenovsky V, Hwang SJ, Lin G, Redfield MM, Borlaug BA. Right heart dysfunction in heart failure with preserved ejection fraction. Eur Heart J. 2014; 35:3452–3462. doi: 10.1093/eurheartj/ehu193.

19. Mankowski RT, Yende S, Angus DC. Long-term impact of sepsis on cardiovascular health. Intensive Care Med. 2019;45(1):78–81. doi:10.1007/s00134-018-5173-1

20. Heart Failure and Sepsis: Cardiologists have developed a new ‘risk profile’ tool to identify heart failure patients at risk of dying from sepsis. Mark Nicholls reports, European Heart Journal, Volume 40, Issue 5, 01 February 2019, Pages 409–410

21. https://www.cfrjournal.com/articles/Right-Ventricular-Failure. as accessed on 9/19/20

22. Biswas A. Right heart failure in acute respiratory distress syndrome: An unappreciated albeit a potential target for intervention in the management of the disease. Indian J Crit Care Med. 2015;19(10):606–609. doi:10.4103/0972-5229.167039

23. Melly L, Torregrossa G, Lee T, Jansens JL, Puskas JD. Fifty years of coronary artery bypass grafting. J Thorac Dis. 2018;10(3):1960–1967. doi:10.21037/jtd.2018.02.43

24. Weiss AJ, Elixhauser A. Trends in Operating Room Procedures in U.S. Hospitals, 2001– 2011: Statistical Brief #171. 2014 Mar. In: Healthcare Cost and Utilization Project (HCUP) Statistical Briefs [Internet]. Rockville (MD): Agency for Healthcare Research and Quality (US); 2006 Feb–. PMID: 24851286.

25. Brown JC, Gerhardt TE, Kwon E. Risk Factors For Coronary Artery Disease. [Updated 2020 Jun 6]. In: StatPearls [Internet]. Treasure Island (FL): StatPearls Publishing; 2020 Jan-. Available from: https://www.ncbi.nlm.nih.gov/books/NBK554410/

26. Papanicolas I, Woskie LR, Jha AK. Health Care Spending in the United States and Other High-Income Countries. JAMA. 2018 Mar 13;319(10):1024-1039. doi: 10.1001/jama.2018.1150. Erratum in: JAMA. 2018 May 1;319(17):1824. PMID: 29536101.

27. https://doi.org/10.1016/j.amjcard.2019.08.038. as accessed on 10/03/20

28. Ong MK, Romano PS, Edgington S, et al. Effectiveness of Remote Patient Monitoring After Discharge of Hospitalized Patients With Heart Failure: The Better Effectiveness After Transition -- Heart Failure (BEAT-HF) Randomized Clinical Trial [published correction appears in JAMA Intern Med. 2016 Apr;176(4):568] [published correction appears in JAMA Intern Med. 2016 Jun 1;176(6):871]. JAMA Intern Med. 2016;176(3):310–318. doi:10.1001/jamainternmed.2015.7712

29. Gupta A, Fonarow GC. The Hospital Readmissions Reduction Program-learning from failure of a healthcare policy. Eur J Heart Fail. 2018;20(8):1169–1174. doi:10.1002/ejhf.1212

30. Psotka MA, Fonarow GC, Allen LA, et al. The Hospital Readmissions Reduction Program: Nationwide Perspectives and Recommendations: A JACC: Heart Failure Position Paper. JACC Heart Fail. 2020;8(1):1–11. doi:10.1016/j.jchf.2019.07.012

31. Bilchick K, Moss T, Welch T, et al. Improving Heart Failure Readmission Costs and Outcomes With a Hospital-to-Home Readmission Intervention Program. Am J Med Qual. 2019;34(2):127–135. doi:10.1177/1062860618788436

32. Zohrabian A, Kapp JM, Simoes EJ. The economic case for US hospitals to revise their approach to heart failure readmission reduction. Ann Transl Med. 2018;6(15):298. doi:10.21037/atm.2018.07.30

33. https://www.ahajournals.org/doi/10.1161/CIR.0000000000000560 as accessed on 9/25/20

34. Amritphale A, Chatterjee R, Chatterjee S, et al. Predictors of 30-Day Unplanned Readmission After Carotid Artery Stenting Using Artificial Intelligence [published online ahead of print, 2021 Apr 9]. Adv Ther. 2021;10.1007/s12325-021-01709-7. doi:10.1007/s12325-021-01709-7

35. Sehgal, Mukul; Custodio, Haidee; Amritphale, Nupur; Davis, Nita; Vidal, Rosa; Amritphale, Amod 1212: Demographics Characteristics and Risk Factors for 30-Day Unplanned Pediatric Sepsis Readmissions, Critical Care Medicine: January 2021 - Volume 49 - Issue 1 - p 609 doi: 10.1097/01.ccm.0000730736.60916.b8

36. Amritphale A, Amritphale N. Refractory Angina: the Current State of Mechanical Therapies. Current Cardiology Reports. 2019;21(6):46.

37. Amritphale Amod, Nupur Amritphale, and Chowdhury H. Ahsan. “Are Biodegradable Third Generation Drug Eluting Stents the Answer to Instent Restenosis?.” Cardiology and Angiology: An International Journal (2014): 15-40.

38. Asad ZUA, Khan SU, Amritphale A, et al. Early vs Late Discharge in Low-Risk STElevation Myocardial Infarction Patients Treated With Percutaneous Coronary Intervention: A Systematic Review and MetaAnalysis. Cardiovascular Revascularization Medicine. 2020;21(11):1360-1368.

39. Amritphale A, Amritphale N, Dubey D. Smartphone Applications Providing Information about Stroke: Are We Missing Stroke Risk Computation Preventive Applications? J Stroke. 2017 Jan;19(1):117. doi: 10.5853/jos.2016.01004r. PMID: 28178403; PMCID: PMC5307936.

40. Dubey D, Amritphale A, Sawhney A, Dubey D, Srivastav N. Analysis of YouTube as a source of information for West Nile Virus infection. Clin Med Res. 2014 Dec;12(3-4):129–32. doi: 10.3121/cmr.2013.1194. Epub 2014 Feb 26. PMID: 24573700; PMCID: PMC4317155.

41. Vadyala S.R., Betgeri S.N., Sherer E.A., Amritphale A.. Prediction of the number of covid-19 confirmed cases based on k-means-lstm. 2020. 2006.14752.

