## Supplementary material for "Causes, Predictors, and Costs of Unplanned 30-Day Readmissions in Patients with Right Heart Failure": Tables and Figures: RHF Tables&Figures.docx

**Table 1:** Baseline characteristics, demographics, comorbidities for readmission analysis of right heart failure.

|  | **No early readmissions**  **(n=5641; 79.0%)** | **30 day readmission**  **(n=1501; 21.0%)** | **Overall**  **(n=7142; 100%)** | **P-Value** |
| --- | --- | --- | --- | --- |
| Age(yrs) (median[IQR]) | 68 [57 – 78] | 67 [56 – 77] | 68 [57 – 78] | 0.019 |
| Female | 2965 (52.6%) | 800 (53.3%) | 3765 (52.7%) | 0.633 |
| Weekend admission | 1156 (20.5%) | 358 (23.9%) | 1514 (21.2%) | 0.005 |
| **Comorbidities** |  |  |  |  |
| Left ventricular systolic dysfunction | 538 (9.5%) | 183 (12.2%) | 721 (10.1%) | 0.002 |
| Left ventricular diastolic dysfunction | 1078 (19.1%) | 341 (22.7%) | 1419 (19.9%) | 0.002 |
| Combined Left ventricular systolic and diastolic dysfunction | 265 (4.7%) | 84 (5.6%) | 349 (4.9%) | 0.151 |
| Biventricular failure | 68 (1.2%) | 15 (1%) | 83 (1.2%) | 0.508 |
| Valvular heart disease | 876 (15.5%) | 224 (14.9%) | 1100 (15.4%) | 0.563 |
| Myocarditis | 594 (10.5%) | 186 (12.4%) | 780 (10.9%) | 0.041 |
| Pericardial diseases | 228 (4%) | 54 (3.6%) | 282 (3.9%) | 0.432 |
| Atrial ﬁbrillation | 2357 (41.8%) | 712 (47.4%) | 3069 (43%) | <0.001 |
| Acute myocardial infarction | 287 (5.1%) | 74 (4.9%) | 361 (5.1%) | 0.804 |
| Prior PCI | 352 (6.2%) | 114 (7.6%) | 466 (6.5%) | 0.059 |
| Prior CABG | 18 (0.3%) | 12 (0.8%) | 30 (0.4%) | 0.011 |
| CAD | 1890 (33.5%) | 557 (37.1%) | 2447 (34.3%) | 0.010 |
| Cardiac dysrhythmias | 2709 (48%) | 785 (52.3%) | 3494 (48.9%) | 0.003 |
| Cardiac arrest | 75 (1.3%) | 9 (0.6%) | 84 (1.2%) | 0.020 |
| Cardiogenic shock | 246 (4.4%) | 44 (2.9%) | 290 (4.1%) | 0.012 |
| Impella support | 13 (0.2%) | 4 (0.3%) | 17 (0.2%) | 0.800 |
| IABP support | 35 (0.6%) | 5 (0.3%) | 40 (0.6%) | 0.185 |
| Vasopressor support | 143 (2.5%) | 44 (2.9%) | 187 (2.6%) | 0.395 |
| Pulmonary hypertension | 3442 (61%) | 898 (59.8%) | 4340 (60.8%) | 0.381 |
| COPD and bronchiectasis | 2295 (40.7%) | 657 (43.8%) | 2952 (41.3%) | 0.031 |
| Pneumonia (except tuberculosis) | 815 (14.4%) | 227 (15.1%) | 1042 (14.6%) | 0.510 |
| Respiratory failure / arrest | 2622 (46.5%) | 577 (38.4%) | 3199 (44.8%) | <0.001 |
| Obesity | 1972 (35%) | 428 (28.5%) | 2400 (33.6%) | <0.001 |
| OSA | 1302 (23.1%) | 319 (21.2%) | 1621 (22.7%) | 0.129 |
| Septicemia | 532 (9.4%) | 144 (9.6%) | 676 (9.5%) | 0.848 |
| Amyloidosis | 9 (0.2%) | 6 (0.4%) | 15 (0.2%) | 0.071 |
| Hematological disorders | 2659 (47.1%) | 851 (56.7%) | 3510 (49.1%) | <0.001 |
| Cancer | 581 (10.3%) | 172 (11.5%) | 753 (10.5%) | 0.194 |
| Diabetes mellitus | 2142 (38%) | 621 (41.3%) | 2763 (38.7%) | 0.017 |
| CKD | 2027 (35.9%) | 739 (49.2%) | 2766 (38.7%) | <0.001 |
| AKI | 1816 (32.2%) | 566 (37.7%) | 2382 (33.4%) | <0.001 |
| Organ Transplant | 55 (1%) | 29 (1.9%) | 84 (1.2%) | 0.002 |
| Hepatic failure | 219 (3.9%) | 52 (3.5%) | 271 (3.8%) | 0.451 |
| Thyroid disorders | 1000 (17.7%) | 307 (20.5%) | 1307 (18.3%) | 0.015 |
| Tobacco abuse | 903 (16%) | 254 (16.9%) | 1157 (16.2%) | 0.400 |
| Alcohol abuse | 227 (4%) | 65 (4.3%) | 292 (4.1%) | 0.599 |
| Dyslipidemia | 2218 (39.3%) | 592 (39.4%) | 2810 (39.4%) | 0.936 |
| HTN | 345 (6.1%) | 83 (5.5%) | 428 (6%) | 0.395 |
| Fluid and electrolyte disorder | 2653 (47%) | 751 (50%) | 3404 (47.7%) | 0.038 |
| **APR DRG Mortality risk** |  |  |  | <0.001 |
| Minor likelihood of dying | 82 (1.5%) | 9 (0.6%) | 91 (1.3%) |  |
| Moderate likelihood of dying | 1588 (28.2%) | 323 (21.5%) | 1911 (26.8%) |  |
| Major likelihood of dying | 2721 (48.2%) | 871 (58%) | 3592 (50.3%) |  |
| Extreme likelihood of dying | 1250 (22.2%) | 299 (19.9%) | 1549 (21.7%) |  |
| **APR DRG Severity of Illness** |  |  |  | <0.001 |
| Minor loss of function (includes cases with no comorbidity or complications) | 46 (0.8%) | 11 (0.7%) | 57 (0.8%) |  |
| Moderate loss of function | 903 (16%) | 170 (11.3%) | 1073 (15%) |  |
| Major loss of function | 3335 (59.1%) | 949 (63.2%) | 4284 (60%) |  |
| Extreme loss of function | 1356 (24%) | 371 (24.7%) | 1727 (24.2%) |  |
| **Primary expected payer** |  |  |  | <0.001 |
| Medicare | 3792 (67.4%) | 1073 (71.6%) | 4865 (68.2%) |  |
| Medicaid | 700 (12.4%) | 218 (14.5%) | 918 (12.9%) |  |
| Private | 851 (15.1%) | 168 (11.2%) | 1019 (14.3%) |  |
| Self-pay | 125 (2.2%) | 29 (1.9%) | 154 (2.2%) |  |
| No charge | 17 (0.3%) | 5 (0.3%) | 22 (0.3%) |  |
| Other | 145 (2.6%) | 6 (0.4%) | 151 (2.1%) |  |
| **Quartile of median household income** |  |  |  | 0.006 |
| 0–25th | 1685 (30.5%) | 517 (34.8%) | 2202 (31.4%) |  |
| 26th –50th | 1578 (28.6%) | 425 (28.6%) | 2003 (28.6%) |  |
| 51st–75th | 1330 (24.1%) | 321 (21.6%) | 1651 (23.6%) |  |
| 76th –100th | 925 (16.8%) | 221 (14.9%) | 1146 (16.4%) |  |
| **Hospital bed size** |  |  |  | 0.398 |
| Small | 937 (16.6%) | 247 (16.4%) | 1184 (16.6%) |  |
| Medium | 1613 (28.6%) | 405 (27%) | 2018 (28.3%) |  |
| Large | 3090 (54.8%) | 850 (56.6%) | 3940 (55.2%) |  |
| **Control/Ownership of Hospital** |  |  |  | 0.715 |
| Government, nonfederal | 625 (11.1%) | 171 (11.4%) | 796 (11.1%) |  |
| Private, not-profit | 4552 (80.7%) | 1216 (81%) | 5768 (80.8%) |  |
| Private, invest-own | 463 (8.2%) | 114 (7.6%) | 577 (8.1%) |  |
| **Hospital Urban Rural Designation** |  |  |  | 0.176 |
| Large metropolitan areas with at least 1 million residents | 3130 (55.5%) | 874 (58.2%) | 4004 (56.1%) |  |
| Small metropolitan areas with less than 1 million residents | 1948 (34.5%) | 495 (33%) | 2443 (34.2%) |  |
| Micropolitan areas | 427 (7.6%) | 95 (6.3%) | 522 (7.3%) |  |
| Not metropolitan or micropolitan (non-urban residual) | 136 (2.4%) | 37 (2.5%) | 173 (2.4%) |  |
| **Hospital teaching status** |  |  |  | 0.150 |
| Metropolitan non-teaching hospital | 1057 (18.7%) | 262 (17.5%) | 1319 (18.5%) |  |
| Metropolitan teaching hospital | 4020 (71.3%) | 1107 (73.8%) | 5127 (71.8%) |  |
| Non-metropolitan hospital | 563 (10%) | 132 (8.8%) | 695 (9.7%) |  |
| **Discharge destination** |  |  |  | <0.001 |
| Home/self-care | 2649 (47%) | 626 (41.7%) | 3275 (45.9%) |  |
| Home health care | 103 (1.8%) | 22 (1.5%) | 125 (1.8%) |  |
| Discharge against medical advice | 66 (1.2%) | 37 (2.5%) | 103 (1.4%) |  |
| **Length of stay and cost analysis** |  |  |  |  |
| Index admission length of stay(days) (median[IQR]) | 5 [3-9] | 6 [3-10] | 5 [3-9] | 0.037 |
| Index admission cost (U.S.$) (median[IQR]) | 12661 [7342 – 23985] | 13873 [7886 – 23729] | 12881 [7439 – 23857] | 0.198 |

**Table 2:** Multivariable logistic regression analysis for predictors of early readmissions.

|  | ***P* value** | **Odds ratio** |
| --- | --- | --- |
| **Comorbidities** |  |  |
| Left ventricular systolic dysfunction | 0.013 | 1.294 (1.055 - 1.588) |
| Left ventricular diastolic dysfunction | 0.000 | 1.325 (1.139 - 1.54) |
| Combined Left ventricular systolic and diastolic dysfunction | 0.588 | 1.079 (0.82 - 1.418) |
| Biventricular failure | 0.483 | 0.812 (0.454 - 1.453) |
| Valvular heart disease | 0.051 | 0.844 (0.712 - 1.001) |
| Myocarditis | 0.882 | 0.985 (0.807 - 1.203) |
| Pericardial diseases | 0.447 | 0.885 (0.645 - 1.213) |
| Atrial ﬁbrillation | 0.014 | 1.417 (1.072 - 1.873) |
| Acute myocardial infarction | 0.576 | 1.083 (0.818 - 1.434) |
| Prior PCI | 0.534 | 1.082 (0.845 - 1.384) |
| Prior CABG | 0.033 | 2.359 (1.071 - 5.197) |
| CAD | 0.318 | 1.075 (0.933 - 1.239) |
| Cardiac dysrhythmias | 0.480 | 0.905 (0.686 - 1.194) |
| Cardiac arrest | 0.082 | 0.529 (0.258 - 1.084) |
| Cardiogenic shock | 0.005 | 0.587 (0.406 - 0.848) |
| Impella support | 0.470 | 1.602 (0.447 - 5.745) |
| IABP support | 0.322 | 0.602 (0.22 - 1.644) |
| Vasopressor support | 0.073 | 1.414 (0.968 - 2.065) |
| Pulmonary hypertension | 0.403 | 0.948 (0.836 - 1.074) |
| COPD and bronchiectasis | 0.000 | 1.358 (1.188 - 1.552) |
| Pneumonia (except tuberculosis) | 0.055 | 1.186 (0.996 - 1.411) |
| Respiratory failure / arrest | 0.000 | 0.71 (0.621 - 0.812) |
| Obesity | 0.000 | 0.686 (0.594 - 0.792) |
| OSA | 0.321 | 0.925 (0.794 - 1.078) |
| Septicemia | 0.757 | 0.967 (0.783 - 1.195) |
| Amyloidosis | 0.191 | 2.055 (0.698 - 6.054) |
| Hematological disorders | 0.000 | 1.341 (1.185 - 1.517) |
| Cancer | 0.206 | 1.131 (0.935 - 1.368) |
| Diabetes mellitus | 0.135 | 1.105 (0.97 - 1.259) |
| CKD | 0.000 | 1.607 (1.402 - 1.843) |
| AKI | 0.052 | 1.142 (0.999 - 1.306) |
| Organ Transplant | 0.133 | 1.448 (0.893 - 2.348) |
| Hepatic failure | 0.162 | 0.791 (0.57 - 1.098) |
| Thyroid disorders | 0.018 | 1.202 (1.033 - 1.398) |
| Tobacco abuse | 0.688 | 1.036 (0.872 - 1.232) |
| Alcohol abuse | 0.827 | 1.034 (0.767 - 1.394) |
| Dyslipidemia | 0.766 | 0.98 (0.86 - 1.117) |
| HTN | 0.066 | 1.277 (0.984 - 1.657) |
| Fluid and electrolyte disorder | 0.694 | 1.025 (0.906 - 1.16) |
| **Primary expected payer** (Reference: Medicaid) |  |  |
| Medicare | 0.979 | 0.997 (0.805 - 1.236) |
| Private | 0.001 | 0.661 (0.521 - 0.839) |
| Self-pay | 0.213 | 0.755 (0.485 - 1.175) |
| No charge | 0.732 | 1.198 (0.428 - 3.354) |
| Other | 0.000 | 0.14 (0.062 - 0.314) |
| **Age groups** (Reference: Age>75years) |  |  |
| 18-44 | 0.000 | 2.511 (1.907 - 3.307) |
| 45-64 | 0.000 | 1.815 (1.502 - 2.193) |
| 65-74 | 0.000 | 1.391 (1.183 - 1.636) |
| **Gender** (Ref: male) |  |  |
| Female | 0.301 | 1.068 (0.943 - 1.21) |
| **Hospital teaching status** (Ref: Non-metropolitan hospital) |  |  |
| Metropolitan non-teaching hospital | 0.312 | 1.085 (0.927 - 1.27) |
| Metropolitan teaching hospital | 0.871 | 1.02 (0.801 - 1.299) |

**Figure 1:** Flowchart describing data extraction with inclusion/exclusion patient encounters

**Patients with unplanned 30 day readmission:**

Un-weighted number: 811

Weighted number*: 1501

**Patients with NO unplanned 30 day readmission:**

Un-weighted number: 3048

Weighted number*: 5640

**Total index admissions:**

Un-weighted number: 6737

**Total index patients meeting inclusion/exclusion criteria:**

Un-weighted number: 3859

Weighted number*: 7142

**Excluded patients:**

(Un-weighted numbers)

Patients with death on index admission: 461

Missing data for death on index discharge: 0

Patients with index discharge in December: 2417

* Weighted numbers: for national estimates as recommended by Health Care Utilization Project (HCUP)

**Figure 2:** Multivariable logistic regression analysis – Forrest Plot

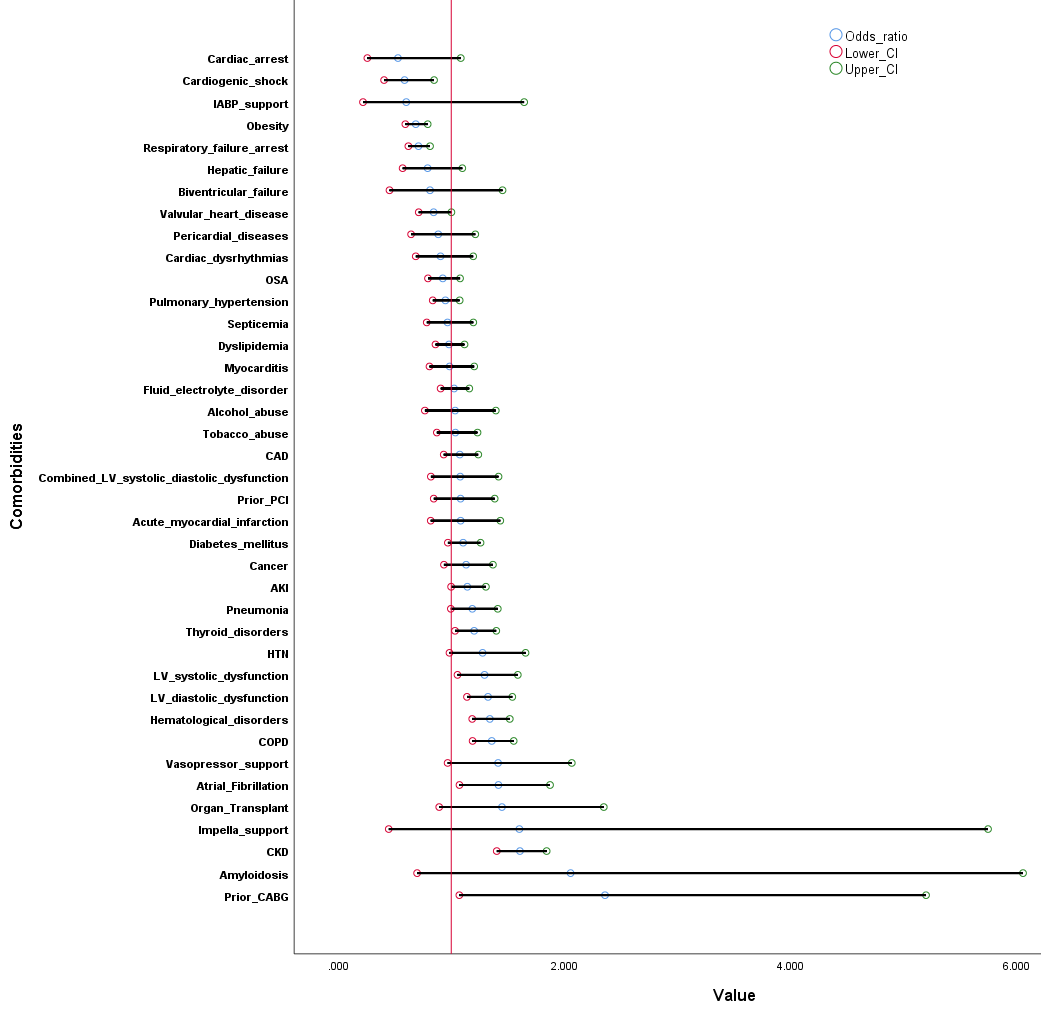

**Figure 3:** Leading primary diagnosis categories for readmissions encounters

|  |  |  |
| --- | --- | --- |
| Heart Failure | 326.749 | 21.8178 |
| Septicemia | 144.976 | 9.68039 |
| Resp Failure | 126.775 | 8.46503 |
| COPD | 54.7274 | 3.65427 |
| Cardiovascular & Cerebrovascular Disorders | 51.1624 | 3.41623 |
| Pneumonia | 48.1113 | 3.2125 |
| Dysrhythmias | 42.3499 | 2.8278 |
| Chronic Kidney Disease | 42.1003 | 2.81113 |
| Acute Kidney Injury | 40.6923 | 2.71712 |
| Gastrointestinal Hemorrhage | 35.154 | 2.34731 |

**Appendix Table 1:** Classification of Clinic Classification Software refined codes (CCS and CCSR) for readmissions Causes and comorbidities.

| **Comorbidities** | **CCSR / ICD-10 CM codes** |
| --- | --- |
| Right heart failure | 'I50810', 'I50811', 'I50812', 'I50813', 'I50814' |
| Left ventricular systolic dysfunction | 'I5020', 'I5021', 'I5022', 'I5023' |
| Left ventricular diastolic dysfunction | 'I5030', 'I5031', 'I5032', 'I5033' |
| Combined Left ventricular systolic and diastolic dysfunction | 'I5040', 'I5041', 'I5042', 'I5043' |
| Biventricular failure | 'I5082' |
| Valvular heart disease | CIR003 |
| Myocarditis | CIR005 |
| Pericardial diseases | CIR006 |
| Atrial ﬁbrillation | I480, I481, I4811, I4819, I482, I4828, I4820, I4821, I4891 |
| Acute myocardial infarction | CIR009 |
| Prior PCI | Z955; Z9861 |
| Prior CABG | I25700; I25701; I25708; I25709; I25710; I25711; I25718; I25719; I25720; I25721; I25728; I25729; I25730; I25731; I25738; I25739; I25790; I25791; I25798; I25799; I25810 |
| CAD | CIR011 |
| Cardiac dysrhythmias | CIR017 |
| Cardiac arrest | CIR018 |
| Cardiogenic shock | R570 |
| Impella support | 5A0211D, 5A0221D |
| IABP support | 5A02110, 5A02210 |
| Vasopressor support | 3E043XZ, 3E030XZ, 3E033XZ, 3E040XZ |
| Pulmonary hypertension | CIR014 |
| COPD and bronchiectasis | RSP008 |
| Pneumonia (except tuberculosis) | RSP002 |
| Respiratory failure / arrest | RSP012 |
| Obesity | END009 |
| OSA | G4733 |
| Septicemia | INF002 |
| Amyloidosis | E852, E853, E854, E858, E8581, E8582, E8589, E859 |
| Hematological disorders | BLD001 TO BLD010 |
| Cancer | NEO001 to NEO074 |
| Diabetes mellitus | END002; END003 |
| CKD | GEN003 |
| AKI | GEN002 |
| Organ Transplant | FAC023 |
| Hepatic failure | DIG018 |
| Thyroid disorders | END001 |
| Tobacco abuse | MBD024 |
| Alcohol abuse | MBD017 |
| Dyslipidemia | END010 |
| HTN | CIR007 |
| Fluid and electrolyte disorder | END011 |
